# Projecting the transition of COVID-19 burden towards the young population while vaccines are rolled out: a modelling study

**DOI:** 10.1101/2021.10.14.21265032

**Authors:** Jun Cai, Juan Yang, Xiaowei Deng, Cheng Peng, Xinhua Chen, Qianhui Wu, Hengcong Liu, Juanjuan Zhang, Wen Zheng, Junyi Zou, Zeyao Zhao, Marco Ajelli, Hongjie Yu

**Affiliations:** Department of Infectious Diseases, Huashan Hospital, School of Public Health, Fudan University; Key Laboratory of Public Health Safety, Ministry of Education, Shanghai, China; Shanghai Institute of Infectious Disease and Biosecurity, Fudan University, Shanghai, China; Laboratory for Computational Epidemiology and Public Health, Department of Epidemiology and Biostatistics, Indiana University School of Public Health, Bloomington, IN, USA

**Author notes:** Corresponding authors: Hongjie Yu, Shanghai Institute of Infectious Disease and Biosecurity, School of Public Health, Fudan University, Shanghai 200032, China. These authors contributed equally to this work (Jun Cai, Juan Yang). These authors are joint senior authors contributed equally to this work (Marco Ajelli, Hongjie Yu).

## Abstract

**Objectives:** SARS-CoV-2 infection causes most cases of severe illness and fatality in older age groups. In China, over 99% of individuals aged ⩾12 years have been fully vaccinated against COVID-19 (albeit with vaccines developed against historical lineages), while 65.0% children aged 3–11 years have been vaccinated their first doses (as of November 12, 2021). Here, we aimed to assess whether, in this vaccination landscape, the importation of Delta variant infections could shift the COVID-19 burden from adults to children.

**Methods:** We developed an age-structured susceptible-infectious-removed model of SARS-CoV-2 transmission dynamics to simulate epidemics triggered by the importation of Delta variant infections and project the age-specific incidence of SARS-CoV-2 infections, cases, hospitalisations, intensive care unit (ICU) admissions, and deaths.

**Results:** In the context of the vaccination programme targeting individuals aged ≥12 years (as it was the case until mid-October 2021), and in the absence of non-pharmaceutical interventions, the importation of Delta variant infections could have led to widespread transmission and substantial disease burden in mainland China, even with vaccination coverage as high as 97% across the eligible age groups. Extending the vaccination roll-out to include children aged 3–11 years (as it was the case since the end of October 2021) is estimated to dramatically decrease the burden of symptomatic infections and hospitalisations within this age group (54% and 81%, respectively, when considering a vaccination coverage of 99%), but would have a low impact on protecting infants (aged 0–2 years).

**Conclusions:** Our findings highlight the importance of including children among the target population and the need to strengthen vaccination efforts by increasing vaccine effectiveness.

## Introduction

The coronavirus disease 2019 (COVID-19) pandemic is still raging worldwide. The highly contagious Delta variant (B.1.617.2) of the severe acute respiratory syndrome coronavirus 2 (SARS-CoV-2) has become the dominant strain across the world^1,2^. The global circulation of the Delta variant has led to a resurgence of COVID-19 cases worldwide, including in areas with high vaccination coverage^3,4^.

A clear positive correlation has been shown between severe illness and fatality from SARS-CoV-2 infection and age^5-7^. COVID-19 vaccines are highly efficacious in preventing severe illness^8-10^. Due to individual choices and age-prioritized vaccination strategies adopted by most countries^11^, vaccine coverage tends to be higher in older adults^12,13^. However, since the summer of 2021, while the Delta variant continued its spread, the number of COVID-19 cases and hospitalisations have trended upwards in many countries with an increasing fraction of children and adolescents^14-17^.

China has been able to avoid the widespread local transmission of SARS-CoV-2 since April 2020^18^. However, the importations of Delta variant infections have caused several local outbreaks, the largest of which occurred in Nanjing city and led to 1,162 reported cases and spilled to 38 other cities^19^. Similar to what was observed in other countries, the Delta surge started in September 2021 in Putian city has been characterized by a high proportion (77/226) of infections in children younger than 12 years^20^.

In China, two inactivated COVID-19 vaccines have already obtained emergency use approval for administration in individuals aged ≥3 years. No vaccine has yet received approval for use in children aged ≤2 years. To prevent the spread of infection in younger age groups, China extended the vaccination programme to include adolescents aged 12–17 years in July 2021, then to include children aged 3-11 years around the end of October 2021 (Figure S2). As of November 12, 2021, 2.24 billion and 84.395 million doses were administered to 12+ years and 3–11 years group, respectively, corresponding to 95% and 32.5% of these two target populations^21^.

The vaccines used in mainland China were developed using historical SARS-CoV-2 lineages and have proven to be highly effective in protecting against severe illness caused by the Delta variant^22-24^. However, their effectiveness in preventing infection appears to be reduced^25^. Moreover, vaccination coverage is highly skewed towards older age groups. It is thus legit to ask whether the spread of Delta variant infections and lack of vaccination in younger age groups could shift the COVID-19 burden towards younger age groups and how this can affect the return to normal. To fill this gap, we developed an age-structured SARS-CoV-2 transmission model to project age-specific epidemiological outcomes, should an epidemic start to unfold. In particular, the model is used to forward simulate alternative vaccination strategies and allows tracking the number of infections, cases, hospitalisations, intensive care unit (ICU) admissions, and deaths by age.

## Methods

### SARS-CoV-2 transmission and vaccination model

We developed an age-structured (16 age groups, Table S2) stochastic compartmental susceptible-infectious-removed (SIR) model to simulate the transmission of SARS-CoV-2 (Delta variant) and assess the health impact of age-targeted vaccination campaigns (Figure S1). Detailed information about the model and adopted parameters are described in Sec. 1.1 of Supplementary Materials and Methods. A summary is presented here.

The model is calibrated to represent the Chinese population and considers the age-mixing patterns quantified prior to the COVID-19 pandemic^26^. Based on contact tracing data in the Hunan province of China^27^, children aged under 15 years were found to have a lower susceptibility to SARS-CoV-2 infection than other age groups, which was confirmed by several independent studies^28^ (Table S1). A sensitivity analysis considering homogeneous susceptibility across age groups is presented in Figure S3. As of November 2021, virtually the entire population of China remains susceptible to SARS-CoV-2 after containment of the first epidemic wave of COVID-19^29,30^, the model is initialized with a fully susceptible population. Simulations were seeded with 40 imported infections^31^ on December 1, 2021 and run forward for 1 year. As sensitivity analyses, we consider also 10 and 20 seeds (Table S1). We considered a basic reproductive number *R*_*0*_ = 6 for the Delta variant^32-34^. It is still unclear whether the Delta variant has a shorter generation time than the original lineage, with studies reporting opposite results: an analysis conducted in Singapore found no significant difference^35^, while a study conducted in UK and one in China estimated a shorter duration than for historical lineages^36,37^. Given this non-conclusive evidence, in the baseline analysis we set the average generation time of the Delta variant to be 7 days, in line with estimates for the original lineages^38^. We then considered a shorter generation time of 4.6 days^39^ in a sensitivity analysis (Table S1).

### Vaccination strategy and vaccine effectiveness

We considered two vaccination strategies: (1) vaccination of individuals aged ≥12 years (here referred to as the “adults+adolescents” vaccination strategy), in agreement with the vaccination programme in place in China until mid-October 2021; (2) as in strategy (1), but the target population was extended to include children aged 3–11 years, as it was the case in China starting from October 28, 2021–this strategy is referred to here as the “adults+adolescents+children” vaccination strategy. We consider that susceptible individuals only are eligible for vaccination, mimicking the programme implemented in China. Booster doses are not considered in this study. Moreover, we accounted for individuals with contraindications against vaccination and pregnant women as they were excluded from the Chinese vaccination programme (Table S2)^39^. We simulated the daily distribution of vaccine doses according to the observed and projected vaccination capacity (see Sec. 1.2 in Supplementary Materials and Methods for details).

We considered a two-dose vaccine with a 21-day interval between doses. Fourteen days after the second dose, the vaccine efficacy in protecting against an infection caused by the Delta variant was set at 54.3%^39^. The considered different vaccine effectiveness (VE) against each of the following clinical endpoints: infection, symptomatic disease, hospitalisation, ICU admission, and death (see Table 1). We further explored a range of higher VE values for sensitivity analyses (Figure S9). We considered VE to be homogeneous across age groups^40-42^. We considered a “leaky” vaccine in which all vaccinated individuals are exposed to a lower risk of infection, which is 1-VE times that of non-vaccinated individuals^43^. We assumed that both the vaccine and natural infection confer protection for longer than the study period (i.e., 1 year)^44^.

**Table 1.**
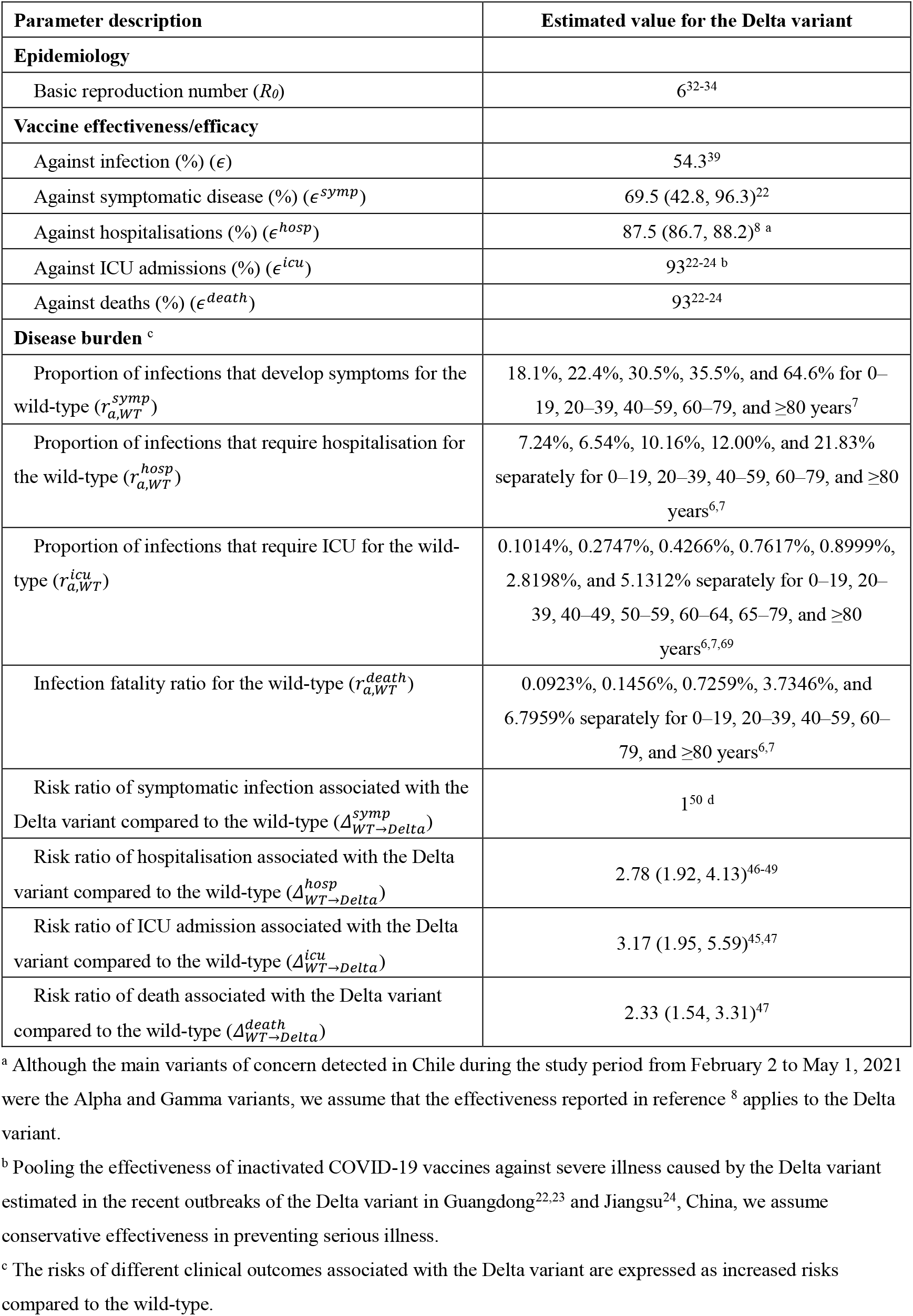
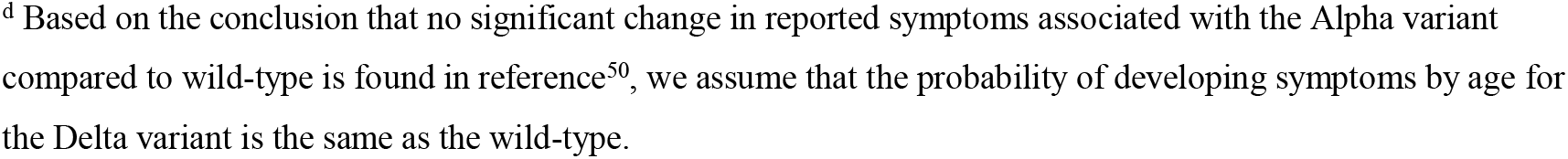
Parameters regulating the transmissibility, vaccine effectiveness/efficacy, and COVID-19 burden.

| Parameter description | Estimated value for the Delta variant |
| --- | --- |
| **Epidemiology** |  |
| Basic reproduction number ($R_0$) | 6<sup>32-34</sup> |
| **Vaccine effectiveness/efficacy** |  |
| Against infection (%) ($\epsilon$) | 54.3<sup>39</sup> |
| Against symptomatic disease (%) ($\epsilon^{symp}$) | 69.5 (42.8, 96.3)<sup>22</sup> |
| Against hospitalisations (%) ($\epsilon^{hosp}$) | 87.5 (86.7, 88.2)<sup>8 a</sup> |
| Against ICU admissions (%) ($\epsilon^{icu}$) | 93<sup>22-24 b</sup> |
| Against deaths (%) ($\epsilon^{death}$) | 93<sup>22-24</sup> |
| **Disease burden <sup>c</sup>** |  |
| Proportion of infections that develop symptoms for the wild-type ($r_{a,WT}^{symp}$) | 18.1%, 22.4%, 30.5%, 35.5%, and 64.6% for 0–19, 20–39, 40–59, 60–79, and $\geq 80$ years<sup>7</sup> |
| Proportion of infections that require hospitalisation for the wild-type ($r_{a,WT}^{hosp}$) | 7.24%, 6.54%, 10.16%, 12.00%, and 21.83% separately for 0–19, 20–39, 40–59, 60–79, and $\geq 80$ years<sup>6,7</sup> |
| Proportion of infections that require ICU for the wild-type ($r_{a,WT}^{icu}$) | 0.1014%, 0.2747%, 0.4266%, 0.7617%, 0.8999%, 2.8198%, and 5.1312% separately for 0–19, 20–39, 40–49, 50–59, 60–64, 65–79, and $\geq 80$ years<sup>6,7,69</sup> |
| Infection fatality ratio for the wild-type ($r_{a,WT}^{death}$) | 0.0923%, 0.1456%, 0.7259%, 3.7346%, and 6.7959% separately for 0–19, 20–39, 40–59, 60–79, and $\geq 80$ years<sup>6,7</sup> |
| Risk ratio of symptomatic infection associated with the Delta variant compared to the wild-type ($\Delta_{WT \rightarrow Delta}^{symp}$) | 1<sup>50 d</sup> |
| Risk ratio of hospitalisation associated with the Delta variant compared to the wild-type ($\Delta_{WT \rightarrow Delta}^{hosp}$) | 2.78 (1.92, 4.13)<sup>46-49</sup> |
| Risk ratio of ICU admission associated with the Delta variant compared to the wild-type ($\Delta_{WT \rightarrow Delta}^{icu}$) | 3.17 (1.95, 5.59)<sup>45,47</sup> |
| Risk ratio of death associated with the Delta variant compared to the wild-type ($\Delta_{WT \rightarrow Delta}^{death}$) | 2.33 (1.54, 3.31)<sup>47</sup> |

<sup>a</sup> Although the main variants of concern detected in Chile during the study period from February 2 to May 1, 2021 were the Alpha and Gamma variants, we assume that the effectiveness reported in reference <sup>8</sup> applies to the Delta variant.
<sup>b</sup> Pooling the effectiveness of inactivated COVID-19 vaccines against severe illness caused by the Delta variant estimated in the recent outbreaks of the Delta variant in Guangdong<sup>22,23</sup> and Jiangsu<sup>24</sup>, China, we assume conservative effectiveness in preventing serious illness.
<sup>c</sup> The risks of different clinical outcomes associated with the Delta variant are expressed as increased risks compared to the wild-type.
<sup>d</sup> Based on the conclusion that no significant change in reported symptoms associated with the Alpha variant compared to wild-type is found in reference<sup>50</sup>, we assume that the probability of developing symptoms by age for the Delta variant is the same as the wild-type.

### Model of COVID-19 burden

The number of infections was produced by the transmission model previously described. To estimate COVID-19 burden, we rescaled the number of infections to obtain estimates of the cumulative incidence of symptomatic cases, hospitalisations, ICU admissions, and deaths under different vaccination strategies using the corresponding age-specific risks estimated for historical lineages and then corrected for the increased severity of the Delta variant (Table 1 – details reported in Sec. 1.3 in Supplementary Materials and Methods). Briefly, by integrating studies conducted in China^45^, Scotland^46^, Canada^47^, and England^48,49^, we estimated that the increased risk associated with the Delta variant was 1.78 (0.92–3.13) for hospitalisation, 2.17 (0.95– 4.59) for ICU admission, and 1.33 (0.54–2.31) for death as compared to the original lineage, respectively (Table 1). We also assumed that the age-specific probability of developing symptoms upon Delta infection is similar to that of the historical lineage. This choice was inspired by the finding that no significant difference was found for the Alpha variant as compared to historical lineages^50^. To compare the severity of disease across different age groups, we calculated the rate ratios as the incidence rate per age group dividing the overall incidence rate for each health outcome under different vaccination strategies.

### Data analysis

For each scenario, 200 stochastic simulations were performed. The outcomes of these simulations determined the distribution of the cumulative number of symptomatic infections, hospitalisations, ICU admissions, and deaths by age. We defined 95% credible intervals as quantiles of the estimated distributions of 0.025 and 0.975.

## Results

### Baseline scenario

The baseline scenario assumes that 40 individuals infected with the Delta variant were introduced in the population on December 1, 2021, and the basic reproduction number *R*_*0*_ was 6 in the absence of interventions and immunity^32-34^. Given vaccination rates data, we project that the “adults+adolescents” vaccination strategy could reach a 97% coverage of the target population (83% of the total population) by November 12, 2021 (Figure 1A).

**Figure 1.**
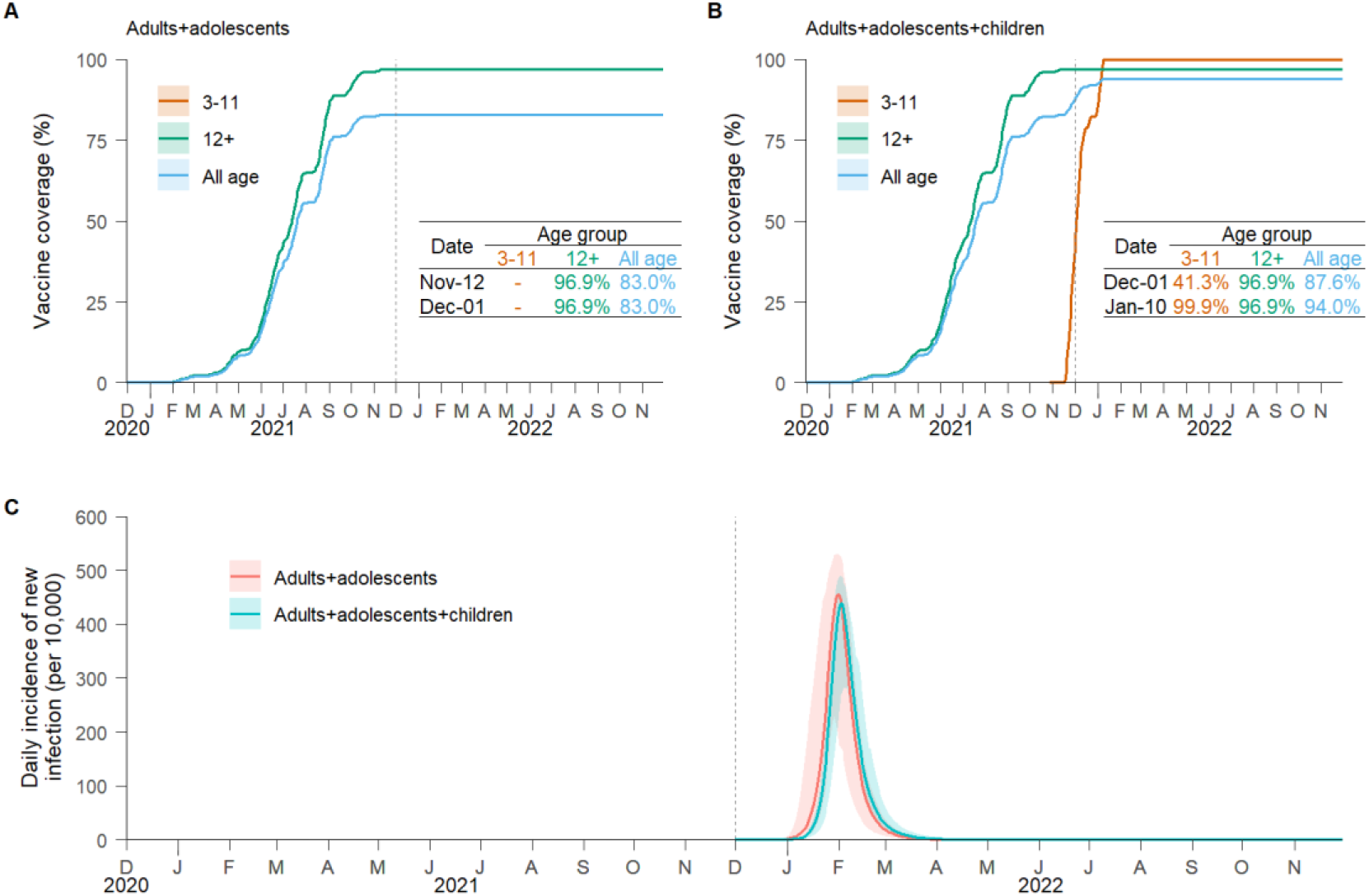
Time series of vaccine coverage and daily incidence of new SARS-CoV-2 Delta variant infections. **A** Age-specific vaccine coverage over time for the “adults+adolescents” vaccination strategy. The vaccination programme was initiated on November 30, 2020 (as first officially reported in China). The vertical dotted lines represent the (simulated) seeding of the infection (December 1, 2021). The subpanel reports a table showing the age-specific coverage. The line corresponds to the mean value, while the shaded area represents the 95% quantile intervals (CI). **B** As A, but for the “adults+adolescents+children” vaccination strategy. **C** Simulated daily incidence of new SARS-CoV-2 Delta variant infections per 10,000 individuals for the two strategies (mean and 95% CI).

Model simulations suggest that the importation of Delta variant infections in December 2021 could have the potential to generate a major epidemic wave in the absence of non-pharmaceutical interventions (NPIs). Such an epidemic is estimated to cause 1,693 (95% CI, 1,596–1,738) symptomatic infections, 827 (95% CI, 757–856) hospitalisations, 39 (95% CI, 37–41) ICU admissions, and 38 (95% CI, 35–39) deaths per 10,000 individuals over 1 year (Figures 1C and 2). These figures correspond to 10–22 fold the disease burden of the initial epidemic in Wuhan, China^6^. Under the adopted vaccination programme, 13% of symptomatic infections and 30% of hospitalisations were estimated to occur in children younger than 12 years (Figure S8), who were ineligible for COVID-19 vaccination until mid-October 2021.

**Figure 2.**
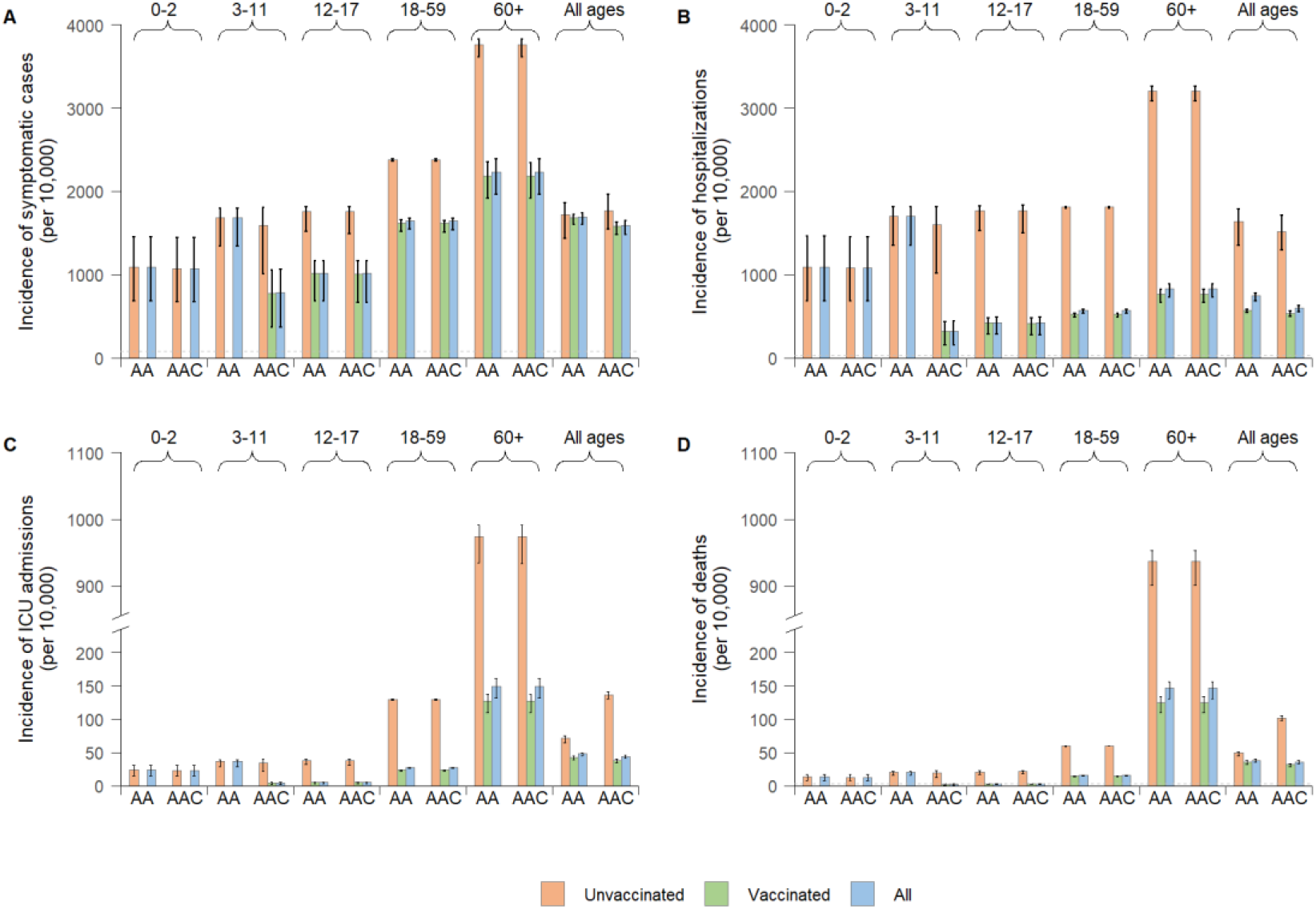
Age profile of projected disease burden caused by an epidemic of imported SARS-CoV-2 Delta variant infections in China. **A** Cumulative number of symptomatic cases per 10,000 individuals after one simulated year by vaccination strategy (AA = “adults+adolescents” vaccination strategy, AAC = “adults+adolescents+children” vaccination strategy), vaccination status, and age group. The vaccinated group are those individuals who are administrated with two doses. **B** As A, but for the incidence of hospitalisations. **C** As A, but for the incidence of ICU admissions. **D** As A, but for mortality. The horizontal dotted lines in **A, B**, and **D** represent the rates of symptomatic cases, hospitalisations, and deaths, respectively, of the first pandemic wave of COVID-19 in Wuhan, China^6^.

Two inactivated COVID-19 vaccines have been licensed for use in children aged 3–17 years^51,52^. However, vaccination has not been implemented among children aged 3–11 years until the end of October 2021. Here, we simulate a scenario where the vaccine is offered to children aged 3–11 years (representing 11.0% of the population) from October 28, 2021 (i.e., the “adults+adolescents+children” vaccination strategy). It is projected that about 41.3% of children aged 3–11 years are fully vaccinated at the (assumed) time of importation of the Delta variant on December 1, 2021 (Figure 1).

As compared to the “adults+adolescents” vaccination strategy, this alternative strategy is estimated to reduce the incidence of symptomatic cases, hospitalisations, ICU admissions, and deaths by 6% (95% CI, 3–12%), 20% (95% CI, 17–27%), 8% (95% CI, 4–15%), and 5% (95% CI, 1–13%), respectively (Figure 2). Despite the beneficial mitigation effect of this strategy, extending vaccination to children is estimated not to be enough to suppress viral circulation. Indeed, 5.1 million deaths (95% CI, 4.7–5.4) (approximately 0.4% of the Chinese population) are projected under this vaccination programme, should NPIs not be implemented (Figure S8).

We used the model to simulate a counterfactual scenario to estimate what would have happened if a vaccination programme had not been implemented. Compared with a no vaccination scenario, the “adults+adolescents” vaccination strategy led to a 47% and 54% increase in the rate ratios of symptomatic infections in children aged 0–2 and 3– 11 years, respectively. We recall that the rate ratio is defined as the incidence rate per age group divided by the overall incidence rate^53^. A higher increase is observed in the rate ratios of hospitalisation for these two population groups: 178% and 191% for children aged 0–2 and 3–11 years, respectively. At the same time, the rate ratios in adults aged ≥60 years is estimated to decrease by 25%. Similar patterns are observed in the rate ratios of ICU admissions and deaths (Figure 3). Extending the vaccination to children aged 3–11 years is projected to dramatically decrease the burden of symptomatic infections and hospitalisations within the same age group by 54% and 81%, respectively (Figure S8). However, due to the strong age-assortativity of contact patterns of this age group (i.e., individuals aged 3–11 years primarily mix with other individuals of the same age) (Figure S4A), extending the vaccination to children does not strongly impact the COVID-19 burden in other age groups (Figure S8). No evident effect is projected on the 0–2-year age group (Figures 3 and S8).

**Figure 3.**
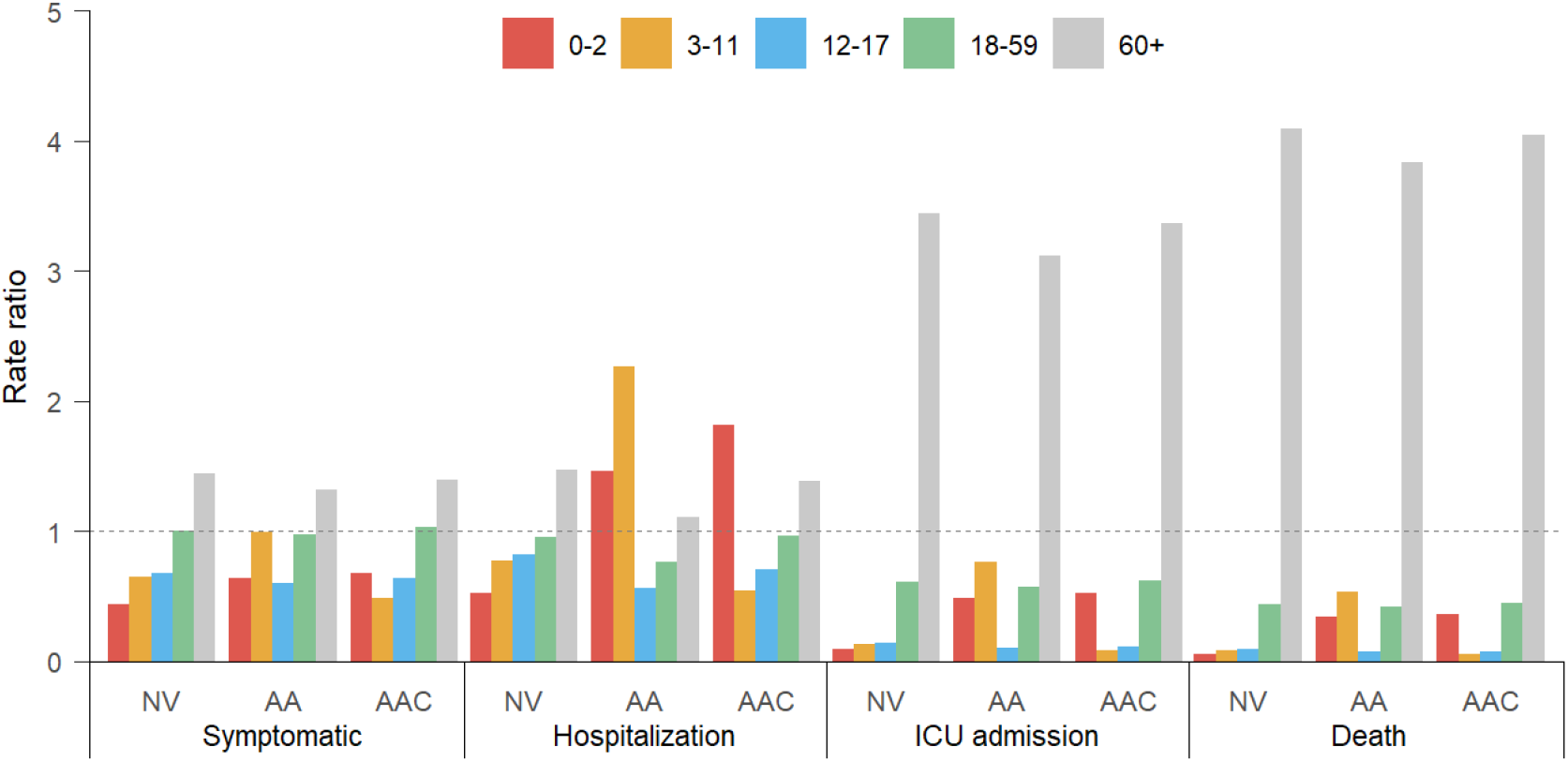
Rate ratios of symptomatic cases, hospitalisations, ICU admissions, and deaths due to SARS-CoV-2 Delta variant infections in China by age group and vaccination strategy. NV = no vaccination, AA = “adults+adolescents” vaccination strategy, AAC = “adults+adolescents+children” vaccination strategy.

Under either the “adults+adolescents” or “adults+adolescents+children” vaccination strategies, the mean incidence of symptomatic cases among unvaccinated individuals is estimated to be 1.5–2.0 fold that of vaccinated individuals, with larger differences observed in terms of the incidence of severe clinical outcomes (3.4–5.0 fold for hospitalisation, 2.9–8.9 fold for ICU admission, and 3.4–8.9 fold for death) (Figure 2).

### Improving VE in the absence/presence of NPIs

In our model, VE in preventing ICU admissions and deaths caused by a Delta variant infection was set at 93%, based on real-world VE studies in Guangdong and Jiangsu, China (Table 1)^22-24^. Should the VE against the infection of the Delta variant be increased to >83% from 54.3% used in the baseline^39^, the “adults+adolescents+children” vaccination strategy is estimated to reduce the number of deaths by 99% even in the absence of any NPIs, leading to <34,000 deaths over 1 year, similar to the annual excess respiratory disease deaths associated with influenza in China^54^ (Figure S9).

We leveraged the developed model to explore the impact of implementing different levels of NPIs on the COVID-19 burden while the “adults+adolescents+children” vaccination strategy is in place and considering different levels of VE against infection. When VE against infection is set at 54.3% (baseline scenario), stringent NPIs capable of maintaining the net reproduction number *R*_*e*_<2.5 are needed, in combination with the vaccination programme, to reduce the yearly COVID-19 death toll to a level similar to that of seasonal influenza^54^ (<44,000 deaths). Should a new vaccine with higher (>75%) VE against the infection be adopted, NPIs could be relaxed. Moderate NPIs able to keep *R*_*e*_<4.4 in combination with the vaccination programme is estimated to be sufficient to decrease the annual number of deaths to less than 60,000 (Figure S9).

### Sensitivity analyses

In the baseline analysis, age-varying susceptibility to SARS-CoV-2 infection was considered. That is, using adults aged 15–64 years as a reference group, children have a 42% lower risk of infection than adults (Table S1)^27^. No estimates are available for susceptibility to Delta variant infections by age. As such, we performed an analysis assuming the same susceptibility to infection across all age groups. The obtained results are consistent with those obtained in the baseline analysis, with average variations lower than 12% (Figure S3).

Another feature able to shape the epidemiology of COVID-19 is the contact pattern of the population. In the baseline analysis, we used pre-pandemic mixing patterns to represent a situation close to the objective of returning to a pre-COVID-19 pandemic lifestyle^26^ (Figure S4A). However, whether mixing patterns will ever return to be the same after the pandemic remains to be seen. As such, we tested the robustness of our findings by considering an alternative contact matrix estimated in Shanghai, China, right after the end of the lockdown in March 2020^55^ (Figure S4B). The obtained results are consistent with those obtained with the pre-pandemic contact matrix, with mean variations in the estimated burden lower than 6% (Figure S5).

We further tested the robustness of our findings by assuming a shorter generation time of 4.6 days and less (10 and 20) imported seed infections. The obtained disease burden is consistent with those obtained in the baseline analysis across age groups and vaccination strategies (Figures S6 and S7).

## Discussion

Our modelling study indicates that, under a vaccination programme targeting individuals aged ≥12 years, symptomatic SARS-CoV-2 infections and hospitalisation would shift towards children and young adolescents in the case of a new COVID-19 wave caused by the Delta variant. These modelling results obtained for China are backed up by empirical evidence from other countries, where an upsurge of COVID-19 cases and hospitalisations among children and adolescents has been reported since the summer of 2021^14,15,53^. Evidence of this epidemiological shift towards children and adolescents comes from the analyses of local outbreaks in Putian city, China, in September 2021, where 34.1% of the reported cases occurred in those aged 0–11 years^20^.

The numbers depicted by the model are related to the relatively low vaccine efficacy against Delta variant infection of the vaccines in use in China (54.3%^39^) as of November 2021. Model projections show that, in the absence of NPIs, even when 97% of the population aged ⩾12 years is fully vaccinated, the importation of Delta variant infections could lead to a new COVID-19 wave and substantial health burden. In particular, the estimated number of deaths over 1 year was 38 per 10,000 individuals as compared, for instance, with 3.6 deaths per 10,000 individuals in the first epidemic wave in Wuhan in early 2020^6^. Extending vaccination to children could mitigate the number of symptomatic cases and severe outcomes but would not be enough to suppress transmission. Relying on NPIs such as border control screenings, border quarantine, isolation, and mask wearing remains key in this phase of widespread circulation of highly transmissible SARS-CoV-2 variants across the globe. This finding is consistent with studies in France, the UK, and the US where, despite the distribution of more effective vaccines (e.g., mRNA vaccines and adenoviral vector vaccines), the transmission is far from being suppressed as of November 2021^56-58^.

To return to normal (i.e., no NPIs), the combination of high vaccine coverage, including children and adolescents, and a highly efficacious vaccine that prevents infection appear to be key to reducing COVID-19 burden to levels closer to those of seasonal influenza. To reach this target, in addition to extending vaccination to children aged 3–11 years as what is being implemented in China since the end of October 2021, our modelling results suggest that vaccine efficacy against the infection would need to be increased to >83%, slightly higher than the efficacy estimated for the mRNA-1273 (Moderna) vaccine^10^. China is currently attempting to develop mRNA vaccines^59^, vaccines that specifically target the Delta variant^60^, multi-valent vaccines, and universal vaccines^61-63^. However, whether such vaccines will become available in the short term is unknown. Moreover, the waning of vaccine-induced immunity may also worsen the situation^57,64-66^. For this reason, several countries recommend booster shots, at least for certain high-risk groups^67,68^. The debate on whether to prioritize the administration of booster doses to older age groups or extend the vaccination programme to those aged 3–11 years merits further modelling investigations. Currently, the best way to protect infants (aged 0–2 years) appears to be by vaccinating their contacts, such as family members.

Although the inactivated vaccines in use in China as of November 2021 show a relatively low effectiveness against mild and moderate disease caused by Delta variant infection, they appear to be highly effective against severe outcomes^8,22-24^. In the context of a constantly changing virus and the evolving epidemiology of the pandemic, it is of great importance to continue to ramp up efforts to increase vaccination coverage by using currently available vaccines. Promoting equitable access to COVID-19 vaccines is critical to decreasing viral circulation and the likelihood of the emergence of new variants of concern.

## Limitations

Our findings should be interpreted by considering the following limitations. First, we assumed that vaccine protection lasted longer than the time of our simulations (1 year). Recent immunological studies have shown that the neutralising antibody titres of inactivated vaccines decline to a low level at 6 months after the second vaccine dose^64,65^, which may indicate waning of vaccine efficacy. However, the correlation with protection from SARS-CoV-2 infection has yet to be proven. Second, we estimated the disease burden potentially caused by the importation of Delta variant infections in the absence or presence of NPIs. The impact of NPIs has been modelled through a simple reduction in transmissibility occurring homogenously with age. We still lack evidence regarding whether this is the case or whether NPIs lead to age-related effects. Moreover, our analysis is not suited to pinpoint which NPIs need to be performed to reach the considered levels of transmission reduction. Further studies exploring this direction are required.

## Conclusions and Public Health Implications

Although vaccination is key to dramatically reducing severe outcomes of SARS-CoV-2 infections and the overall burden of the COVID-19 pandemic, in the absence of NPIs, the importation of Delta variant infections could lead to widespread transmission and substantial disease burden in mainland China, even with vaccination coverage as high as 97% of those aged ≥12 years (VE=54.1% in preventing the infection). Moreover, vaccination programmes targeting individuals aged ≥12 years only can potentially shift the COVID-19 burden towards younger ages. Extending vaccination to children aged 3-11 years would mitigate the disease burden within this age group, but would not be enough to suppress transmission.

Our findings highlight the need to strengthen vaccination efforts by simultaneously extending the target age groups of the vaccination campaign, and elevating VE via booster vaccination and/or developing new highly efficacious vaccines in China.

## Data Availability

The data and code that support the findings of this study will be made available on GitHub upon the acceptance of this manuscript.

## Contributors

H.Y. conceived, designed, and supervised the study. J.C. and X.D. developed the model. J.C., J.Y., X.D., X.C., Q.W., H.L., J.J.Z., W.Z., J.Y.Z., and Z.Z. collected the data. J.C., J.Y., X.D., M.A., and H.Y. interpreted the results. J.C., X.D., and C.P. prepared the tables and figures. J.C. and J.Y. wrote the first draft of the manuscript. M.A. and H.Y. critically revised the content. All authors approved the final manuscript as submitted and agree to be accountable for all aspects of the work.

## Acknowledgments

This study was supported by grants from the Key Program of the National Natural Science Foundation of China (82130093) and the National Institute for Health Research (NIHR) (grant no. 16/137/109) using UK aid from the UK Government to support global health research. The views expressed in this publication are those of the author(s) and not necessarily those of the NIHR or the UK Department of Health and Social Care.

## Conflicts of interest

H.Y. received research funding from Sanofi Pasteur, GlaxoSmithKline, Yichang HEC Changjiang Pharmaceutical Company, Shanghai Roche Pharmaceutical Company, and SINOVAC Biotech Ltd. M.A. received research funding from Seqirus. Except for research funding from SINOVAC Biotech Ltd, which is related to the data analysis of clinical trials of immunogenicity and safety of CoronaVac, the others are not related to COVID-19. All the other authors have no competing interests.

## Human participant protection

This study collected only publicly available data and did not involve human participants. No institutional review board approval was needed.

