## Supplementary Information for "Projecting the transition of COVID-19 burden towards the young population while vaccines are rolled out: a modelling study"

### 1. Materials and methods

#### 1.1. SARS-CoV-2 transmission and vaccination model

We developed an age-structured stochastic susceptible-infectious-removed (SIR) model to simulate SARS-CoV-2 transmission and vaccination. The Chinese population was divided into 16 age groups, as presented in Table S2. We considered a baseline scenario, where:

- We used age-mixing patterns specific to China that were quantified during the pre-pandemic period[1].
- We assumed an age-dependent susceptibility to SARS-CoV-2 Delta infection that was lower in children under 15 years and higher in adults aged 65+ years (Table S1)[2].
- Asymptomatic and symptomatic individuals are assumed to be equally infectious, and infectiousness is assumed to be the same across age groups[2].
- A two-dose vaccination is assumed to reduce an individual's susceptibility to SAR-CoV-2 infection. We consider the vaccine effectiveness (VE) against SARS-CoV-2 Delta infection to be homogeneous across age groups[3-5].
- As vaccine breakthrough cases increase[6,7], we consider a "leaky" vaccine where all vaccinated individuals are exposed to a lower risk of infection, which is 1-VE times that of non-vaccinated individuals[8].
- We assumed that vaccine- or natural infection-induced protection lasts longer than the period considered (1 year)[9].

To test the impact of age-mixing patterns on the transmission of imported Delta variant infections, a sensitivity analysis considering an alternative contact matrix for China estimated in March 2020 (post-lockdown period)[10] was explored. In addition, homogeneous susceptibility to infection across all age groups was explored as a sensitivity analysis (see Sec. 2.1).

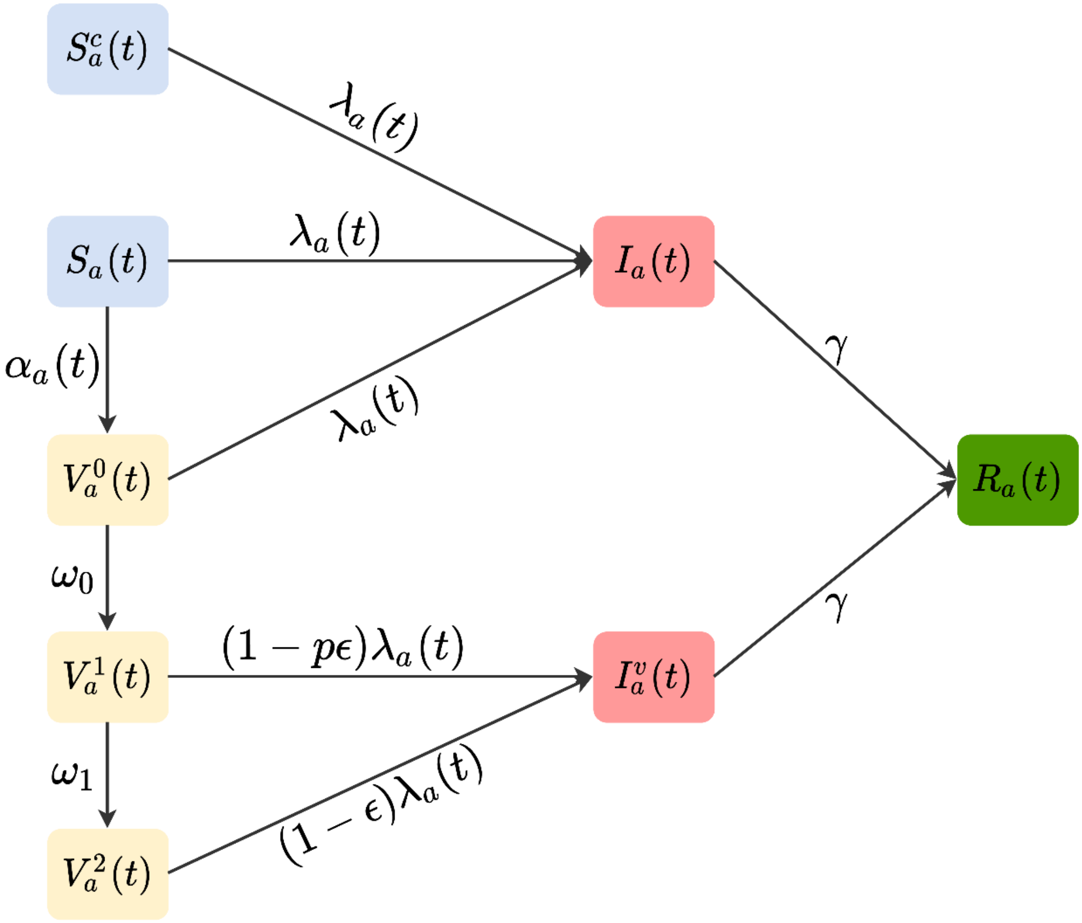

**Figure S1. Schematic representation of SARS-CoV-2 transmission and vaccination model.** Taking the age group $a \in [1, 16]$ at time $t$ as an example, model compartments are defined as follows: unvaccinated susceptible individuals who are ineligible for vaccination due to contraindications and pregnancy $S_a^c(t)$, unvaccinated susceptible individuals who are eligible for vaccination $S_a(t)$, unvaccinated infected individuals $I_a(t)$, removed individuals $R_a(t)$, unprotected individuals who received their first dose within 21 days before time $t$ $V_a^0(t)$, partially vaccinated individuals who received their second dose within 14 days before time $t$ $V_a^1(t)$, vaccinated individuals who received their second dose at least 14 days before time $t$ $V_a^2(t)$, and vaccinated infected individuals $I_a^v(t)$. Note that vaccinated individuals are defined as those administered two doses. The model parameters include: time- and age-dependent force of infection $\lambda_a(t)$, probability of administration of the first dose ($\alpha_a(t)$), recovery rate from infection $\gamma$, relative vaccine efficacy against infection right after administration of the second dose $p$ as compared with maximum protection (i.e., after ramp-up of the second dose) $\epsilon$, time interval between the administration of the first and second dose $(1/\omega_0)$ and maximum protection $(1/\omega_1)$.

The SARS-CoV-2 transmission and vaccination model is schematically represented in Figure S1 and described by the following system of differential equations:

$$
\begin{aligned}
\frac{dS_a^c(t)}{dt} &= -\lambda_a(t)S_a^c(t),\\
\frac{dS_a(t)}{dt} &= -\lambda_a(t)S_a(t) - \alpha_a(t)S_a(t),\\
\frac{dI_a(t)}{dt} &= \lambda_a(t)\big(S_a^c(t) + S_a(t) + V_a^0(t)\big) - \gamma I_a(t),\\
\frac{dV_a^0(t)}{dt} &= \alpha_a(t)S_a(t) - \lambda_a(t)V_a^0(t) - \omega_0 V_a^0(t),\\
\frac{dV_a^1(t)}{dt} &= \omega_0 V_a^0(t) - (1 - p\epsilon)\lambda_a(t)V_a^1(t) - \omega_1 V_a^1(t),\\
\frac{dV_a^2(t)}{dt} &= \omega_1 V_a^1(t) - (1 - \epsilon)\lambda_a(t)V_a^2(t),\\
\frac{dI_a^v(t)}{dt} &= (1 - p\epsilon)\lambda_a(t)V_a^1(t) + (1 - \epsilon)\lambda_a(t)V_a^2(t) - \gamma I_a^v(t),\\
\frac{dR_a(t)}{dt} &= \gamma\big(I_a(t) + I_a^v(t)\big),
\end{aligned}
\tag{1}
$$

where:

- $S_a^c$ represents the number of unvaccinated susceptible individuals who are ineligible for vaccination due to contraindications and pregnancy in age group $a$ (Table S2).
- $S_a$ represents the number of unvaccinated susceptible individuals who are eligible for vaccination in age group $a$ (Table S2).
- $I_a$ represents the number of unvaccinated infected individuals of age group $a$.
- $R_a$ represents the number of recovered or removed individuals of age group $a$.
- $V_a^0$ represents the number of individuals administered the first dose in age group $a$. We assumed that the second dose was administered 21 days after the first dose, namely $1/\omega_0 = 21$ days.
- $V_a^1$ represents the number of individuals in age group $a$ who were administered their second dose within 14 days. The interval between administration and full protection of the second dose was 14 days, namely $1/\omega_1 = 14$ days.

- $V_a^2$ represents the number of individuals of age group $a$ who received their second dose for at least 14 days.
- $I_a^v$ represents the number of vaccinated infected individuals of age group $a$.
- $\epsilon$ represents the expected VE against infection after ramp-up of the second dose.
- $p$ represents the relative VE against infection right after the administration of the second dose as compared with the maximum protection $\epsilon$.

Susceptible individuals of age group $a$ at time $t$ are exposed to a time- and age-dependent force of infection $\lambda_a(t)$, which is defined as:

$$\lambda_a(t) = (1-\varphi)\beta r_a \sum_{\tilde{a}} C_{a,\tilde{a}} \frac{I_{\tilde{a}} + I_{\tilde{a}}^v}{N_{\tilde{a}}}, \tag{2}$$

where:

- $\beta$ is a scaling factor shaping SARS-CoV-2 transmissibility in the absence of non-pharmaceutical interventions (NPIs) such as face masks or hand hygiene precautions, computed by assuming basic reproduction number $R_0$ = 6[11-13], as estimated for the SARS-CoV-2 Delta variant.
- $\varphi$ is a coefficient representing the reduction in transmissibility due to NPIs.
- $r_a$ is the relative susceptibility to SARS-CoV-2 infection in age group $a$ (Table S1).
- $C_{a,\tilde{a}}$ represents the age-group-specific contact matrix, whose elements describe the mean number of daily contacts that an individual in age group $a$ has with individuals in age group $\tilde{a}$ (Figure S4).
- $N_{\tilde{a}}$ represents the number of individuals in age group $\tilde{a}$ (Table S2).

Given the value of $R_0 = 6$, the distribution of the age-specific susceptibility profile ($r_a$), and the distribution of the bootstrapped contact matrix $C_{a,\tilde{a}}$, the distribution of transmission rate $\beta$ is calculated analytically through the equation described in our previous work[14]. When considering a set of NPIs that are capable of bringing the net reproduction number to a value $R_e < R_0$, we rescale the transmission rate $\beta$ by a factor

$(1-\varphi)$, where $\varphi = 1 - R_e/R_0$. We tested a range value of $R_e$ = 1.1~5.9 with a step of 0.2 to explore the impact of different intensities of NPIs adopted on the disease burden from Delta variant infections in China.

Given that vaccinated and unvaccinated individuals infected with the Delta variant have similar proliferation and peak Ct values[15-17], it appears that there is a limited difference in infectiousness despite vaccination. Thus, we assumed that all infectious compartments despite vaccination status ($I_a$ and $I_a^v$) have the same average duration of infectiousness ($1/\gamma$), which corresponds to the length of generation time in an SIR model[18]. We considered an average generation time of 7 days[19] in the main analysis. While studies also show that the Delta variant transmits more quickly than previously circulating variants[20,21], a shorter generation time of 4.6 days is considered within sensitivity analyses (see Sec. 2.4).

We simulated the daily vaccination process in China as follows. First, we prioritised those individuals who need to receive their second dose at time $t$; then, the remaining doses are randomly administered to those who are eligible to receive their first dose. At each time $t$, the first dose is administered to a fraction $\alpha_a(t)$ of unvaccinated susceptible individuals in age group $a$:

$$\alpha_a(t) = \frac{d_a(t)}{S_a(t)}, \quad (3)$$

where $d_a(t)$ represents the number of first doses to be administered to individuals in age group $a$ at time $t$ under the designed vaccination strategies.

We assumed that the first dose does not confer protection[22,23], that partially vaccinated individuals who were administered their second dose within 14 days are protected by a proportion $p$ of the expected vaccine efficacy $\epsilon$ (i.e., $p\epsilon$), and that fully vaccinated individuals who were administered their second dose after at least 14 days are protected by the expected vaccine efficacy $\epsilon$ (see Sec. 1.4). The proportion $p$ = 0.84 (42.5%/50.7%) was estimated from the efficacies of CoronaVac vaccine reported in the PROFISCOV study[4]. The CoronaVac vaccine is one of inactivated vaccines widely used in the ongoing mass vaccination programme in China.

Simulation results were obtained using a stochastic version of the model described above with a time step of 0.25 days. A total of 200 simulations were run for each scenario, sampling at each simulation a different value from the joint distribution of transmission rate $\beta$, the bootstrapped contact matrices $C_{a,\tilde{a}}$ and the age-specific susceptibility profile $r_a$.

#### 1.2. Daily vaccination capacity projection

As described in our previous work[24], we projected the daily vaccination capacity based on the cumulative doses of COVID-19 vaccines administered in China released by the National Health Commission (NHC) until November 12, 2021[25]. The first data on cumulative doses administered in China was released on November 30, 2020. Until March 22, 2021, the NHC began to regularly report the cumulative number of doses administered in China per day. Before March 22, the daily vaccination capacity was interpolated by assuming a constant daily vaccination rate between two adjacent available data points.

Based on the cumulative doses (3.53 millions) allocated to children aged 3-11 years as of October 29, 2021[26] and the doses (3.332 millions) administered at October 28[25], 2021, we assumed that the roll-out started to include children aged 3-11 years at October 28, 2021. As the daily vaccination capacity reported between October 28 and November 12, 2021 also included booster doses, we obtained the daily doses administered to children aged 3-11 years during the same period by multiplying a proportion (84.395/118.461), where the numerator is the cumulative doses (in millions) administered to children aged 3-11 years as of November 12, 2021[27], and the denominator is the total doses (in millions) administered in China between October 28 and November 12, 2021.

For projecting the daily vaccination capacity forwards from November 12 to December 31, 2021, we then fit a quadratic polynomial to meet the following three criteria: 1) the total doses administered between October 28 and December 31, 2021 should fulfil the NHC's target that the vaccination of children aged 3-11 years will be completed by the end of December 2021[27]; 2) the daily vaccination rate at December 31, 2021 is assumed to be as low as observed at October 1, 2021. 3) the curve starts at November 12, 2021. The projected daily doses administrated to children aged 3-11 years are shown in Figure S2. The daily vaccination capacity since 2022 are assumed to be the same as the doses administered at January 1, 2022, which is predicted by the fitted curve.

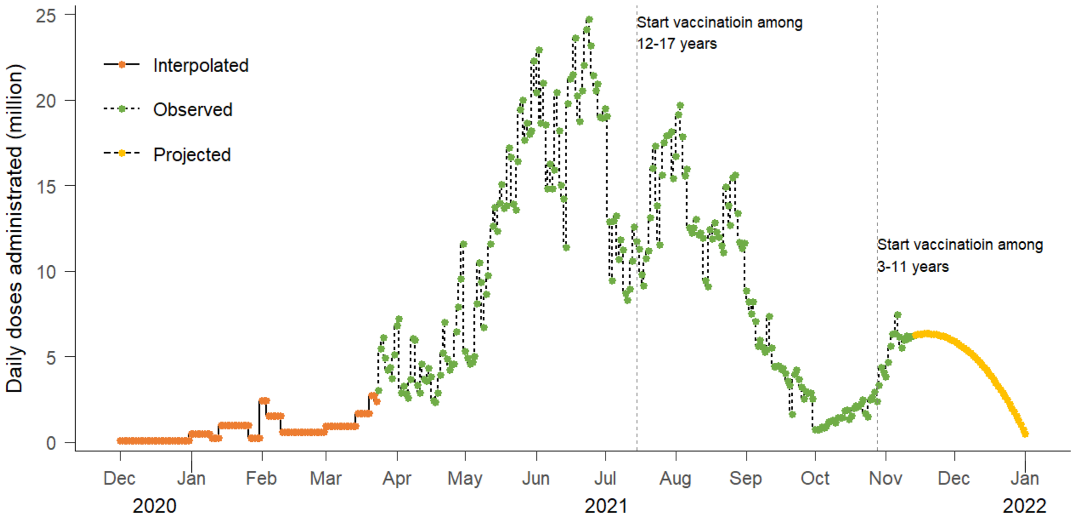

**Figure S2. Daily number of vaccine doses administered in China.** The daily doses administered in China between March 22 and November 12, 2021, were obtained from the National Health Commission. After November 12, 2021, the daily doses administered were projected, whereas before March 22, 2021, the daily doses administered were interpolated. Note that the curve after October 28, 2021 represents the daily doses administered to children aged 3-11 years.

#### 1.3. COVID-19 burden model

The main model outputs are the age-specific number of new infections per day in unvaccinated and vaccinated individuals $i_{\mathrm{a}}(t)$ and $i_a^v(t), t = 1, 2, \cdots, 365$. Mathematically, these are computed as follows:

$$i_a(t) = I_a(t+1) - (1-\gamma)I_a(t), \quad (4)$$

$$i_a^v(t) = I_a^v(t+1) - (1-\gamma)I_a^v(t). \quad (5)$$

The COVID-19 burden of each vaccination scenario was evaluated in terms of the cumulative incidence of symptomatic cases, hospitalisations, ICU admissions, and deaths over the 1 year $B^{type}$, $type \in \{symp, hosp, icu, death\}$, with:

$$B^{type} = \sum_{t=1}^{365}\sum_{a=1}^{16}\left(r_a^{type} i_{\mathrm{a}}(t) + r_a^{v\_type} i_a^v(t)\right), \quad (6)$$

where $r_a^{type}$ represents the risks of developing symptoms, being hospitalized, being admitted to ICUs, and dying among unvaccinated persons infected the Delta variant in age group $a$; $r_a^{v\_type}$ represents the corresponding age-specific risks among vaccinated persons infected with the Delta variant. $r_a^{type}$ (see equation 7) and $r_a^{v\_type}$ (see equation 8 and 9) are estimated in following equations (7-9). Note that we are interested in the total incidence of these epidemiological outcomes. Therefore, we did not account for delays between infection and different clinical outcomes (i.e., symptomatic cases, hospitalisations, ICU admissions, and deaths).

Scarce information on the risks of progressing from infection to different clinical outcomes for the Delta variant has been reported in China because several local outbreaks caused by imported Delta cases were quickly contained. Meanwhile, an increasing number of studies suggest that the Delta variant is associated with a greater risk of adverse outcomes, including hospitalisation, ICU admission, and death for patients, than the wild-type or Alpha variants[28-31]. Therefore, we estimated the age-specific risk $r_a^{type}$, $type \in \{symp, hosp, icu, death\}$ using the following equation:

$$r_a^{type} = r_{a,WT}^{type} \times \Delta_{WT\rightarrow Delta}^{type}, \quad (7)$$

where $r_{a,WT}^{type}$ represents the risks of developing symptoms, being hospitalized, being admitted to ICUs, and dying among those infected with the wild-type strain in age group $a$. $\Delta_{WT\to Delta}^{type}$ represents the (adjusted) risk ratio (or hazard ratio) of the corresponding clinical outcomes caused by the Delta variant compared with the wild-type variant.

The age-specific risks $r_{a,WT}^{type}$ for the wild-type infection are presented in Table 1, which were estimated as described in our previous work[14]. By pooling different studies from China[28], Scotland[29], Canada[31] and England[30,32], we estimated the risk ratio associated with the Delta variant as $\Delta_{WT\to Delta}^{hosp}$= 2.78 (1.92–4.13) for hospitalisation, $\Delta_{WT\to Delta}^{icu}$= 3.17 (1.95–5.59) for ICU admission, and $\Delta_{WT\to Delta}^{death}$= 2.33 (1.54–3.31) for death compared to the wild-type strain, respectively. However, we assumed $\Delta_{WT\to Delta}^{symp}$= 1, inspired by the finding that there was no significant change in the symptoms reported by those with the Alpha or wild-type variants[33] (Table 1). We estimated $\Delta_{WT\to Delta}^{hosp}$ by linking the risk ratio of hospitalisation associated with the Delta variant compared to the Alpha variant, and the risk ratio of hospitalisation associated with the Alpha variant compared to the wild-type strain.

Reduced risks of clinical outcomes are observed for vaccinated infections compared to unvaccinated infections[34]. We adopted below method[35] to adjust the risks of progressing from infection to different clinical outcomes for vaccinated individuals, $r_a^{v\_type}$, with:

$$r_a^{v\_type} = r_a^{type} \times \left(1 - \epsilon^{type|infect}\right), \quad (8)$$

$$\epsilon^{type} = \epsilon + (1-\epsilon) \times \epsilon^{type|infect}, \quad (9)$$

where $\epsilon^{type|infect}$ represents the VE in preventing different clinical outcomes conditionally on infection with the Delta variant; while $\epsilon^{type}$ represents the overall

VE in preventing symptomatic infection, hospitalisation, ICU admission, and death (Table 1; see Sec. 1.4 for details).

#### 1.4. Estimation of vaccine effectiveness/efficacy

We estimated the overall effectiveness/efficacy of the inactivated COVID-19 vaccine against different clinical endpoints (i.e., infection, symptomatic cases, hospitalisation, ICU admission, and death) caused by the Delta variant. Following the work by Khoury et al.[36], we predicted vaccine protection against the Delta variant infection ($\epsilon$=54.3%) with the efficacy against the wild-type infection for BBIBP-CorV vaccine (73.5%)[3], as well as a reduction in the fold change of neutralising antibodies for the Delta variant from *in vitro* neutralisation assay (2.4-fold, 95% CI: 1.1–5.2)[37]. Details on the method for predicting vaccine protection against the Delta variant are given in Chen et al.[38].

The effectiveness of full vaccination against symptomatic COVID-19 caused by the Delta variant ($\epsilon^{symp}$=69.5%, 95% CI: 42.8–96.3%) is obtained from a real-world study on the outbreak of the Delta variant in Guangdong, China[22,23].

The effectiveness of the inactivated Chinese COVID-19 vaccine against hospitalisations caused by the SARS-CoV-2 variants of concern (VOCs) has only been reported in a real-world study in Chile[39]. However, the Alpha and Gamma variants were the main VOCs detected in Chile during the study period (February 2 to May 1, 2021). Therefore, we assumed that the inactivated vaccines used in China are as effective at preventing hospitalisation associated with the Delta variant as the Alpha and Gamma variant (i.e., $\epsilon^{icu}$=87.5%, 95% CI: 86.7–88.2%).

The effectiveness of the inactivated Chinese COVID-19 vaccine against severe illness caused by the Delta variant has also been reported in three studies in China[22,23,40]. However, the effectiveness (100%) estimated from the outbreak of the Delta variant in Guangdong, China, might be overestimated due to the small sample size (only two severe cases who were unvaccinated in the case group)[22,23]. This estimate also contradicts the fact that severe illness among vaccinated individuals was reported in the subsequent Delta outbreak (July-August, 2021) in Jiangsu, China[40]. Thus, we assume a conservative effectiveness (93%) in preventing serious illness, including ICU admission and death, by pooling these estimates from the two outbreaks (i.e., $\epsilon^{icu} = \epsilon^{death}$= 93%).

Considering that the VE against the Delta variant infections might be improved in the future, we explored a range value of $\hat{\epsilon}$ = 50%~95% with a step of 5%. When calculating the COVID-19 burden of these scenarios, the VE against different clinical endpoints, $\hat{\epsilon}^{type}$, $type \in \{symp, hosp, icu, death\}$, were adjusted using the following equation:

$$\hat{\epsilon}^{type} = \max\left(k \times \epsilon^{type}, \epsilon_0^{type}\right) \tag{10}$$

where $k = \hat{\epsilon}/\epsilon$, represents the fold change in VE against infections. $\epsilon_0^{symp}$ = 96.4%, represents the reported highest VE against symptomatic COVID-19 caused by SARS-CoV-2 variants[41], whereas $\epsilon_0^{hosp} = \epsilon_0^{icu} = \epsilon_0^{death}$ = 100%, respectively represents the reported highest VE against hospitalisation, ICU admission, and death.

A summary of the model parameters is presented in Table 1 in the main text, as well as in Tables S1 and S2.

**Table S1. Summary of key parameters used in the model and sensitivity analyses**

| Parameter | Description | Value (or range) | Sensitivity analysis |
|---|---|---|---|
| **Demographic** | | | |
| $N_i$ | Population size for age group *i* | See Table S2 [42] | - |
| $C_{a,\tilde{a}}$ | Age-group-specific contact matrix | Contact matrix in Shanghai before the pandemic (Figure S4A) [1] | Contact matrix for Shanghai in post-lockdown period (Figure S4B) [10] |
| **Epidemiology** | | | |
| $1/\gamma$ | Generation time (days) | 7 [19] | 4.6[21] |
| $r_a$ | Relative susceptibility to SARS-CoV-2 infection at age *a* | $r_a = 0.58$ (95% CI 0.34–0.98) when $a < 15$; $r_a = 1$ for $15 \leq a < 65$; $r_a = 1.65$ (95% CI 1.03–2.65) when $a \geq 65$ [2] | $r_a = 1$ for all age groups (homogeneous susceptibility) |
| $n_0$ | Initial seed infectors | 40 [14] | 10, 20 |
| **Vaccination** | | | |
| $1/\omega_0$ | Interval between the administration of the first dose and second dose (days) | 21 [3] | - |
| $1/\omega_1$ | Delay between the administration of the second dose and achievement of the expected vaccine efficacy (days) | 14 [3] | - |
| $p$ | Relative vaccine efficacy between 0 and $1/\omega_1$ days after the administration of the second dose compared to the expected vaccine efficacy | 0.84 [4] | - |

**Table S2. Overall population, proportion of those with contraindications to vaccination, and pregnant women by age group in China**

| ID | Age group | Population | Contraindications (%) | Pregnant women (%) |
|---|---|---|---|---|
| 1 | 0–2 | 47,208,873 | 0.2 | 0.0 |
| 2 | 3–11 | 158,390,745 | 0.1 | 0.0 |
| 3 | 12–17 | 98,365,257 | 0.1 | 0.3 |
| 4 | 18–24 | 120,465,522 | 0.2 | 4.0 |
| 5 | 25–29 | 97,989,003 | 0.2 | 10.3 |
| 6 | 30–34 | 128,738,970 | 0.3 | 5.2 |
| 7 | 35–39 | 100,091,455 | 0.5 | 2.6 |
| 8 | 40–44 | 96,274,146 | 0.7 | 0.7 |
| 9 | 45–49 | 119,837,617 | 0.9 | 0.3 |
| 10 | 50–54 | 123,445,382 | 1.2 | 0.0 |
| 11 | 55–59 | 98,740,491 | 1.7 | 0.0 |
| 12 | 60–64 | 77,514,139 | 2.2 | 0.0 |
| 13 | 65–69 | 74,149,766 | 2.7 | 0.0 |
| 14 | 70–74 | 44,949,689 | 3.2 | 0.0 |
| 15 | 75–79 | 26,544,616 | 3.2 | 0.0 |
| 16 | ≥80 | 26,618,103 | 2.6 | 0.0 |

### 2. Sensitivity analyses

#### 2.1. Homogeneous susceptibility to infection by age

We evaluated the model sensitivity to the assumption that susceptibility to infection is reduced in children and increased in older adults. In fact, while this may have been the case for historical lineages[2,43], it may not necessarily be correct for the Delta variant. Under an alternative assumption of an equal susceptibility to infection across all age groups, we recalibrated the value of $\beta$ and applied all other conditions as in the baseline scenario. We projected that the disease burden decreased slightly, ranging from 2% to 3% for symptomatic cases and hospitalisations, and about 12% for ICU admissions and death as compared to the baseline, for the "adults+adolescents+children" vaccination strategy (Figure S3).

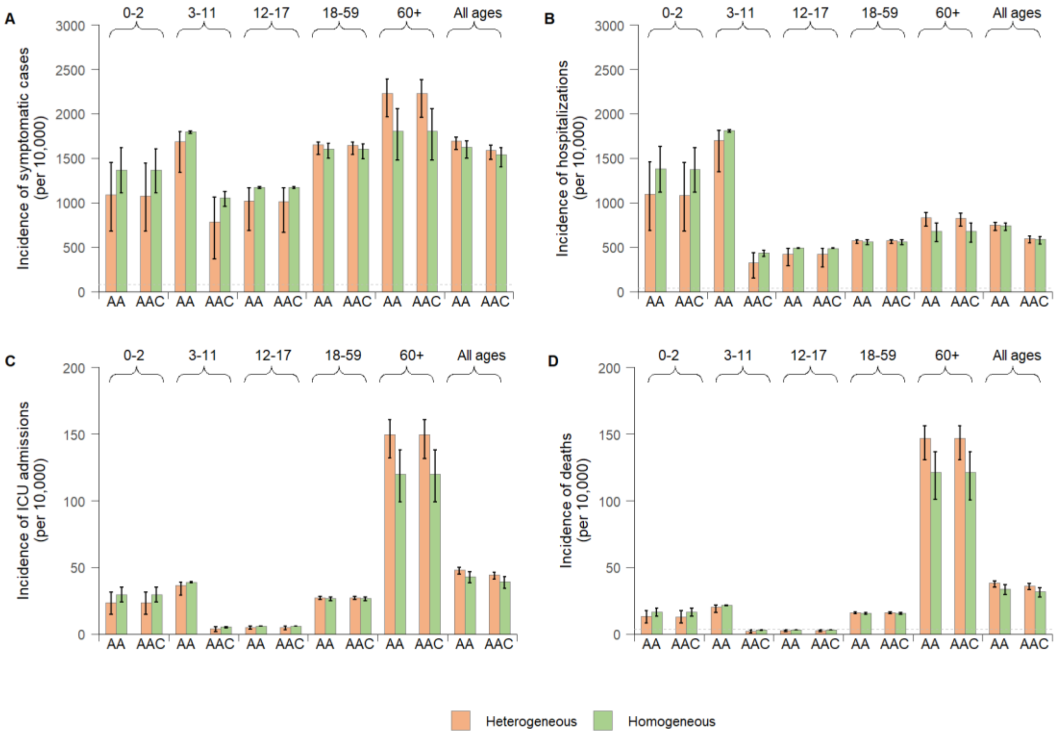

**Figure S3. Disease burden due to SARS-CoV-2 Delta variant infections in China assuming homogeneous or heterogeneous susceptibility to infection. A** Cumulative number of symptomatic cases per 10,000 individuals after one simulated year by vaccination strategy (AA = "adults+adolescents" vaccination strategy, AAC = "adults+adolescents+children" vaccination strategy) and age group for heterogeneous (baseline) and homogeneous susceptibility to infection by age. **B** As A, but for the incidence of hospitalisations. **C** As A, but for the incidence of ICU admissions. **D** As A, but for the incidence of deaths. The horizontal dotted lines in **A**, **B**, and **D** respectively represent the rates of symptomatic cases, hospitalisations, and deaths of the first pandemic wave of COVID-19 in Wuhan, China[44].

### 2.2. Contact patterns in the post-lockdown period

We adjusted the age groups of the contact matrix according to the age groups classified in the mass vaccination campaign in China. Age-mixing patterns specific to Shanghai, China in the pre-pandemic period were used in the main analysis[1] (Figure S4A). As the physical distancing policies adopted during the pandemic have changed the population contact patterns[10,45], we evaluated the model sensitivity to contact patterns derived from different periods. Using an alternative age-mixing pattern in Shanghai, China, estimated in March 2020 (post-lockdown period)[10] (Figure S4B), we recalibrated the value of $\beta$ and applied all other conditions as in the baseline scenario. We projected a 3–6% increase in disease burden in terms of symptomatic cases, hospitalisations, ICU admissions, and deaths, compared to the baseline under the "adults+adolescents+children" vaccination strategy (Figure S5).

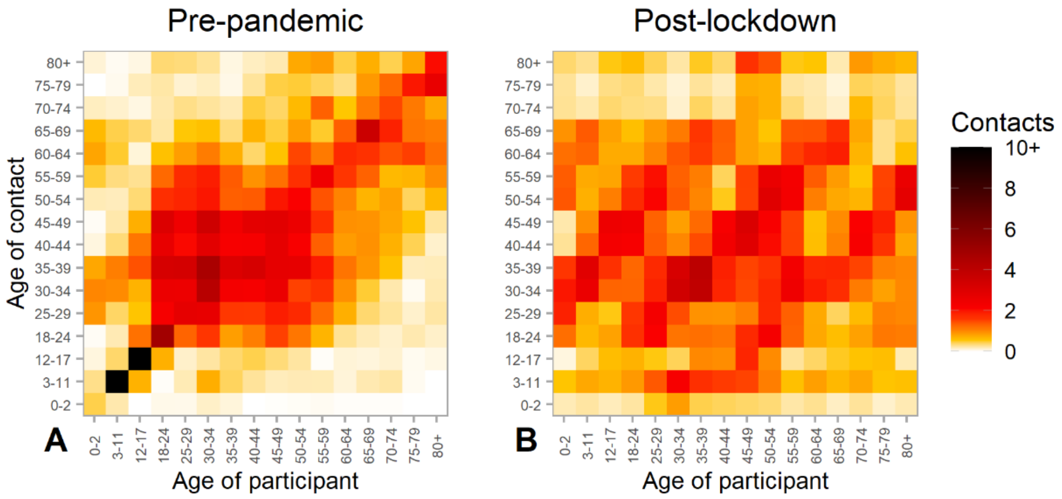

**Figure S4. Age-mixing patterns in China. A** Pre-pandemic contact matrix, which is used in the main analysis. Mixing patterns refer to Shanghai, China in 2017/2018[1]. Each cell of the matrix represents the mean number of daily contacts that an individual in a given age group has with other individuals, stratified by age group. The colour intensity represents the number of daily contacts. **B**

As A, but for the post-lockdown contact matrix. Mixing patterns refer to Shanghai, China, in March 2020, when interventions were relaxed after the lockdown[10].

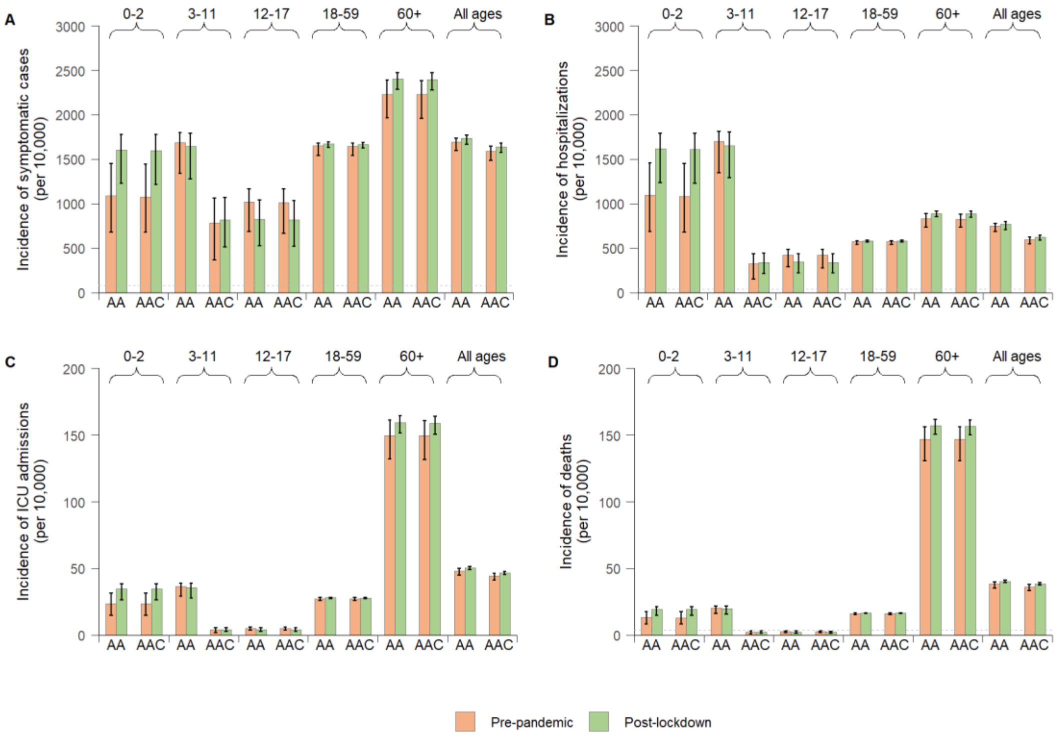

**Figure S5. Disease burden due to SARS-CoV-2 Delta variant infections in China using age-mixing patterns estimated in the post-lockdown period. A** Cumulative number of symptomatic cases per 10,000 individuals after one simulated year by vaccination strategy (AA = "adults+adolescents" vaccination strategy, AAC = "adults+adolescents+children" vaccination strategy) and age group for pre-pandemic (baseline) and post-lockdown contact patterns. **B** As A, but for the incidence of hospitalisations. **C** As A, but for the incidence of ICU admissions. **D** As A, but for the incidence of deaths. The horizontal dotted lines in **A**, **B**, and **D** respectively represent the rates of symptomatic cases, hospitalisations, and deaths of the first pandemic wave of COVID-19 in Wuhan, China[44].

### 2.3. Number of initial seed infectors

In the main analysis, 40 imported infections are seeded to trigger an epidemic. Under China's dynamic zero-case policy, it’s more likely a small number of imported infections that would trigger local outbreaks. Thus, we performed a sensitivity analysis on the number of initial seed infectors and repeated the baseline simulations with 10 and 20 seeding imported infections (Table S1). The resulting disease burden are robust to the number of initial seed infectors across vaccination strategies and age groups (Figure S6).

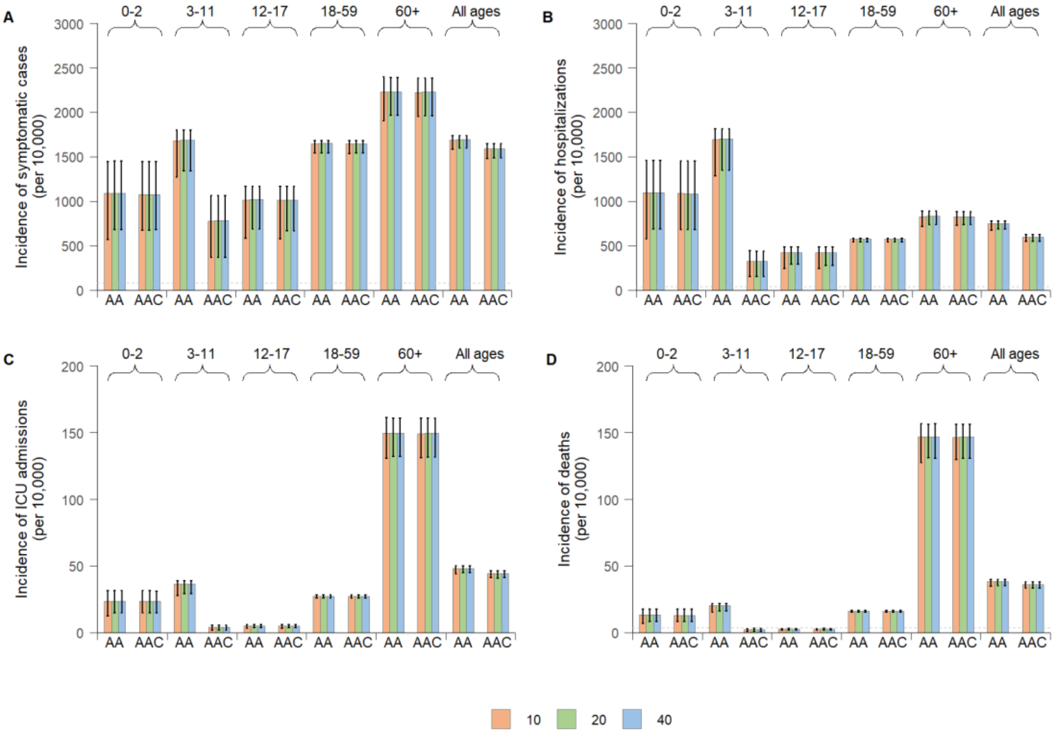

**Figure S6. Disease burden due to SARS-CoV-2 Delta variant infections in China assuming varying number of initial seed infectors. A** Cumulative number of symptomatic cases per 10,000 individuals after one simulated year by vaccination strategy (AA = "adults+adolescents" vaccination strategy, AAC = "adults+adolescents+children" vaccination strategy) and age group for heterogeneous (baseline) and homogeneous susceptibility. **B** As A, but for the incidence of hospitalisations. **C** As A, but for the incidence of ICU admissions. **D** As A, but for the incidence of deaths. The horizontal dotted lines in **A**, **B**, and **D** respectively represent the rates of symptomatic cases, hospitalisations, and deaths of the first pandemic wave of COVID-19 in Wuhan, China[44].

### 2.4. Generation time

We evaluated the model sensitivity to the assumption that the Delta variant has the same generation time ($1/\gamma = 7$ days) as the original lineage[19,46]. However, studies also show that the Delta variant transmits more quickly than previously circulating variants[20,21]. Assuming a shorter generation time of 4.6 days (Table S1), we recalibrated the value of $\beta$ and applied all other conditions as in the baseline scenario. The resulting disease burden are insensitive to generation time across vaccination strategies and age groups (Figure S7). Actually, the incidence of infections is determined by the reproduction number, which remains unaltered under an alternative generation time.

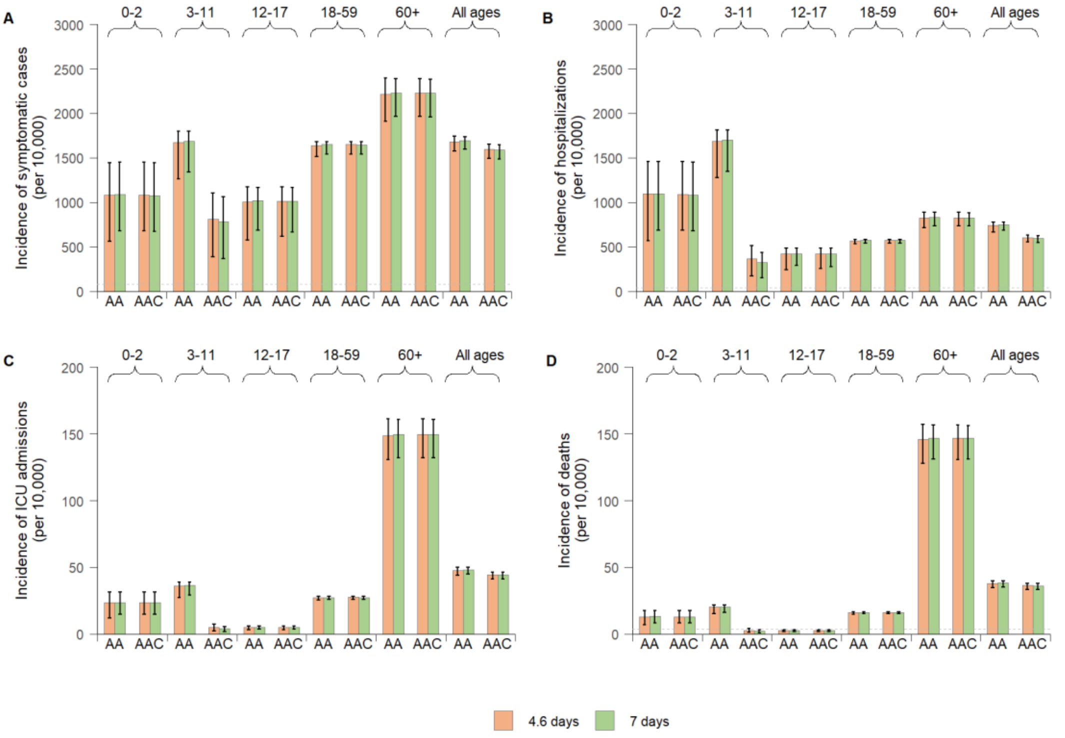

**Figure S7. Disease burden due to SARS-CoV-2 Delta variant infections in China assuming alternative generation time. A** Cumulative number of symptomatic cases per 10,000 individuals after one simulated year by vaccination strategy (AA = "adults+adolescents" vaccination strategy, AAC = "adults+adolescents+children" vaccination strategy) and age group for heterogeneous (baseline) and homogeneous susceptibility. **B** As A, but for the incidence of hospitalisations. **C** As A, but for the incidence of ICU admissions. **D** As A, but for the incidence of deaths. The horizontal dotted lines in **A**, **B**, and **D** respectively represent the rates of symptomatic cases, hospitalisations, and deaths of the first pandemic wave of COVID-19 in Wuhan, China[44].

### 3. Additional results

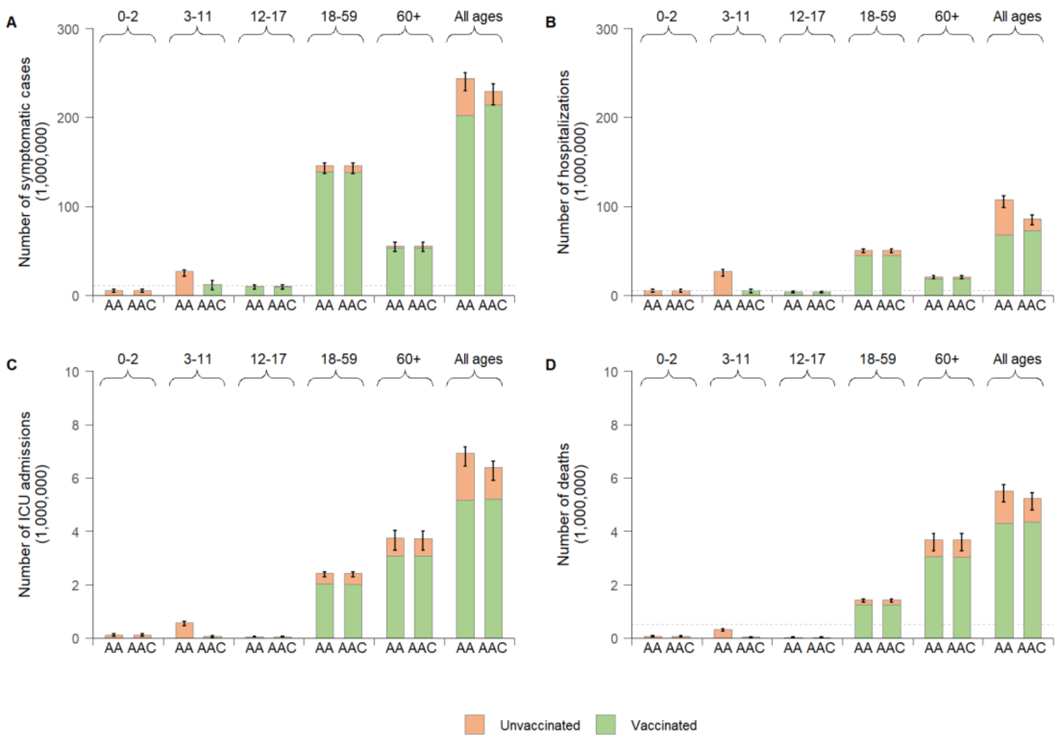

**Figure S8. Cumulative number of symptomatic cases, hospitalisations, ICU admissions, and deaths due to SARS-CoV-2 Delta variant infections in China under the baseline scenario. A** Cumulative number of symptomatic cases after one simulated year by vaccination strategy (AA = "adults+adolescents" vaccination strategy, AAC = "adults+adolescents+children" vaccination strategy), vaccination status, and age group. The vaccinated group are those individuals who have been administrated two doses. **B** As A, but for the number of hospitalisations. **C** As A, but for the number of ICU admissions. **D** As A, but for the number of deaths. The horizontal dotted lines in **A**, **B**, and **D** respectively represent the number of symptomatic cases, hospitalisations, and deaths that would occur in China based on the corresponding rates of the first pandemic wave of COVID-19 in Wuhan, China[44].

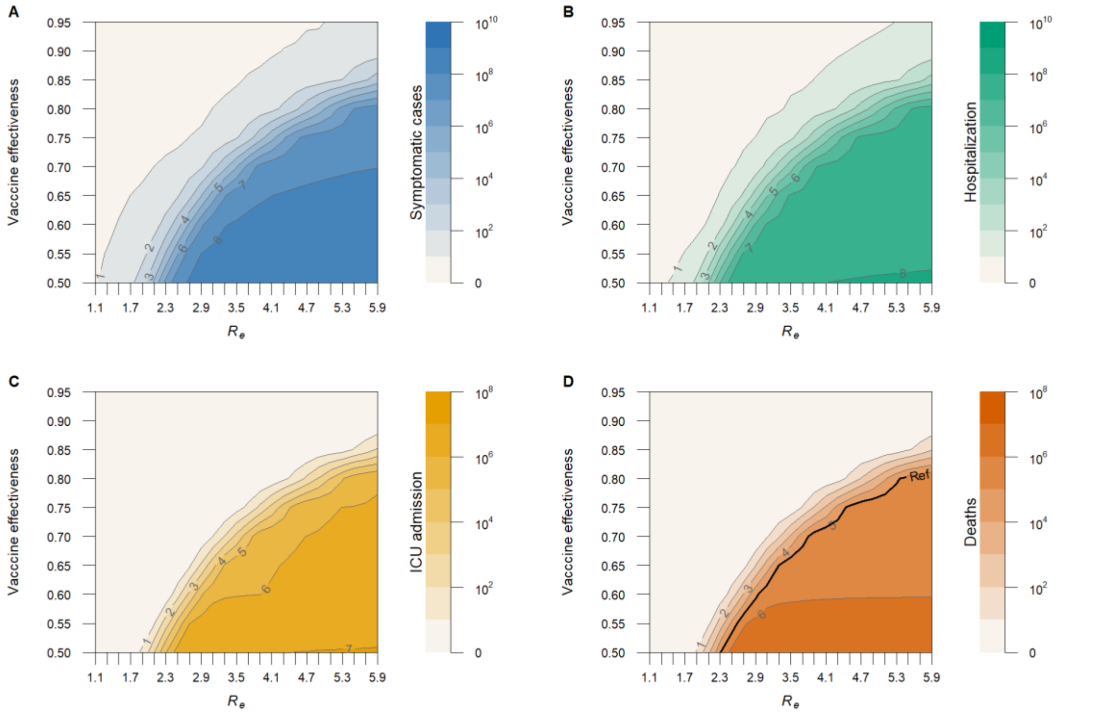

**Figure S9. Impact of adopting NPIs ($R_e$) and increasing vaccine effectiveness (VE) against infections on disease burden due to SARS-CoV-2 Delta variant infections in China under the "adults+adolescents+children" vaccination strategy. A** Cumulative number of symptomatic cases (on a logarithmic scale) after one simulated year as a function of $R_e$ and VE. **B** As A, but for the cumulative number of hospitalisations. **C** As A, but for the cumulative number of ICU admissions. **D** As A, but for the cumulative number of deaths. The solid black reference line in **D** indicates the annual mean of 88,100 influenza-associated excess respiratory deaths in China[42].
